# Descriptive analysis of social determinant of mental health factors in an ethnically diverse Black adult population

**DOI:** 10.1101/2021.10.27.21265590

**Authors:** Aderonke Bamgbose Pederson, Devan Hawkins, Lynette Lartey

## Abstract

**Background:** Black adults are often treated as a homogeneous group in research and health care despite the diversity within the Black population across ethnicity. This study considers ethnicity in assessing the heterogeneity among Black adults across multiple social determinants of mental health. Understanding the diversity within the Black population will help close the gap in mental health disparities by offering a more streamlined approach to meeting unmet mental health needs.

**Methods:** A cross-sectional descriptive study and analysis was conducted among Black adults in the United States (n = 269, ages 18-65) from diverse ethnic backgrounds (African-Americans, African immigrants, Afro-Caribbean immigrants). We calculated mean differences according to ethnicity, citizenship status, age group, and gender in the areas of medical mistrust, use of mental health services, depression symptom severity, mental health knowledge and stigma behavior.

**Results:** African Americans with moderate to severe depression symptoms had greater stigma behavior (mean = 12.2, SD = 3.2), than African Americans who screened in the minimal to mild depression range (mean = 13.1, SD = 3.5). Black immigrants across the spectrum of depression scores had greater stigma than African Americans (p = 0.037). Participants who identified as male had a prevalence of mild depression (5-9) that was 1.7 times higher than those who identified as female. Whereas, those who identified as female had a prevalence that was 1.2 times higher for moderate to severe depression (10-19) and 4.7 times higher for severe depression (20-27) compared to males (p = 0.021). Non-United States citizens reported higher medical mistrust (mean value difference = 0.16) compared to United States citizens (p = 0.011). We found statistically significant differences in depression symptom severity based on gender, prevalence of depression, medical mistrust and stigma behavior across demographic factors within the sample.

**Conclusion:** This study shows key variations across social determinant of mental health factors within the Black adult population. There is a need to better understand the heterogeneity within the Black population in order to improve the effectiveness of programs that seek to reduce mental health disparities.

## Introduction

The majority of studies that assess the needs of Black people focus on comparisons across race with White people and non-Hispanic white people, few studies consider the diversity within the Black population in assessing mental health needs (1, 2). The very few studies that do address the diversity within the Black population in the United States tend to predominantly focus on differences between African Americans and Afro-Caribbean immigrants, leaving out African immigrants (2-4). The limited studies that address differences in mental health needs within the Black population focus on differences in mental health outcomes (such as prevalence of disease) (2, 5), rather than a more in depth approach to variations within the population that influences specific mental health behavior such as social determinants related to mental health knowledge, stigma towards mental illness, help seeking behavior and medical mistrust (6-8). Understanding and addressing these social determinants of mental health by exploring key differences within the Black population provides new ways to reduce mental health disparities (9, 10). While the problem of disparities in mental health outcomes across racial groups (such as differences between white and black counterparts) is evident, the variations within the Black adult population around mental health and mental health behaviors remains unclear and understudied (2, 3, 11).

This study considers ethnicity in assessing the heterogeneity among Black adults across multiple social determinants of mental health in relation to mental health knowledge, stigma behavior (or discrimination), use of mental health services and medical mistrust. A previous study by Williams and colleagues (National Survey of American Life) showed that the prevalence of Major Depressive Disorder (MDD) varied across race and ethnicity and laid a foundation for the importance of understanding the heterogeneity within the Black population (2). While differences in MDD prevalence is relevant clinically, there remains a gap in the literature across differences in psychosocial factors that impact mental health. Our study fills a clinical and research gap in the literature by further describing differences in depression symptom severity across ethnicity and other demographic factors as well as differences among Black adults across psychosocial dimensions of mental health knowledge, stigma behavior, use of mental health services and medical mistrust. No other study specifically assesses the differences among these critical areas of the social determinants of mental health across a sample of African Americans and Black immigrants. This study provides data that lays a foundation towards further examination of within group differences as a means to better address mental health disparities in the Black adult population.

This sample is unique in that we report across various identities of Black adults including across ethnicity (African Americans and Black immigrants), citizenship status, age, and gender. These demographic and social characteristics are considered important in assessment of mental health and mental health needs based on the intersectionality framework (12, 13). Intersectionality refers to the varied identities one may hold, and when applied to health refers to understanding the role of social characteristics may explain health inequities (12, 13). Based on these demographic and social identities, we aimed to examine the mean differences across psychosocial determinants of mental health while considering depression severity. First, we report on ethnicity and depression severity as well as mean outcomes in the area of mental health knowledge, stigma behavior, and self-report willingness to use mental health services for both personal or emotional problems and suicidal ideation. We also report on the distribution of depression severity and medical mistrust across ethnicity, citizenship status, age and gender. We hypothesized that there would be within group variations beyond prevalence of depression that are necessary to consider in our approach to alleviating mental health inequities across the intersectional identities held by Black adults.

## Methods

### Participants, study design and setting

We collected self-report data using an online cross-sectional survey from Black adults (n = 269) residing in the United States from September 2020 to October 2020. Participants included Black adults who identified as African-American, African immigrant, and Afro-Caribbean immigrant. Recruitment occurred in partnership with community based organizations (World Relief Chicago, the United African Organization, Refugee One) using online flyers and posters on social media (email, twitter and Facebook platforms). The eligibility criteria included: 1) identifying as Black, African-American, African or Afro-Caribbean; 2) aged 18-65; 3) currently residing in the United States; and 4) English speaking. Informed consent was completed by each participant prior to participation in study activities. A total of 269 Black adults completed the full survey out of 296 people who submitted a survey (90.9% completion rate).

### Ethical Review

Data collection occurred after obtaining approval by the Institutional Review Board at XXXX (IRB ID #: STU00213136).

### Procedures

Funding for this study was provided by the National Center for Advancing Translational Sciences (NCATS). Convenience sampling was used to collect survey data. The survey included several domains including demographic questionnaire, assessment of mental health knowledge, self-report help-seeking behavior, medical mistrust, mental health stigma behavior and depression screening outcomes. In this manuscript, we report on the diversity across African Americans and Black immigrants based on descriptive analysis of the data collected. Participants received $25 compensation for their time.

### Measures

The measures used were as far as possible validated and standardized instruments. In addition, a demographic questionnaire was used. The following instruments and their psychometric properties are described below.

Demographic Questionnaire: We developed a demographic questionnaire to assess age, gender, race, ethnicity, marital status, education, employment status, income and insurance status. We also assessed residency status (length of stay in the U.S., and birthplace).

The Mental Health Knowledge Schedule (MAKS) (14): The MAKS is composed of two sections, we used the first section to assess general mental health knowledge. We assessed mental health knowledge based on six questions about knowledge about help-seeking, recognition of illness, support methods, employment, treatment and recovery related to mental illness. The second section reviews specific knowledge about common mental health conditions (depression, stress, schizophrenia, bipolar disorder, drug addiction, grief) by assessment of each condition, given our interest in general mental health knowledge, we only used the first section. The first six items are scored on an ordinal scale (1 to 5), higher total scores represent greater knowledge. For this study, we reviewed differences in mental health knowledge among participants based on the first six items. The overall test-retest reliability was moderate to substantial (0.71). The cronbach’s alpha overall internal consistency for items 1 to 6 was 0.65 (moderate) (14).

Reported and Intended Behavior Scale (RIBS) (15): The RIBS measures the reported social distance from people with a mental health problem in the past, current and future time, we used the measure of stigma behavior in the future. The RIBS measures intended future stigma behavior as a continuous variable, based on a total score across four questions with high scores representing less stigmatizing behavior. Responses to the stigma behavior measure include 6 items measured on a Likert scale (*agree strongly* to *disagree strongly* and the option to answer *don’t know*). Each question begins with “in the future,” I would be willing to ‘live’, ‘work’, ‘be a neighbor to’ or ‘have a close friendship’ with someone with a mental health problem. The Cronbach’s alpha was 0.85 for the subscale on stigma behavior. Overall test-retest reliability was 0.75 (15).

The General Help-Seeking Questionnaire (GHSQ) (16): The GHSQ was designed to assess intentions to seek help from several sources (family, friends, and mental health providers, among other sources) and for different problems (personal or emotional problem and suicidal ideation). The cronbach’s alpha for the overall scale is 0.85. The measure includes items on a 7-point scale (1-extemely unlikely, 7-extremely likely) in reference to intentions to seek help. We report on the intention to seek help from a mental health professional (mental health provider, counselor, psychologist or social worker) for a personal or emotional problem and for suicidal ideation.

The depression screening questionnaire, patient health questionnaire (PHQ-9): The PHQ-9 is a 9-item screening measure for depression. The score ranges from 0 to 27 with each of the 9 items scored from 0 (*not at all*) to 3 (*nearly every day*) and higher scores representing greater symptoms of depression (17). PHQ-9 scores represents depressive symptoms that are minimal (0-4), mild (5-9), moderate (10-14), moderate to severe (15-19) and severe (20-27).

The group based medical mistrust scale (GBMMS) (18): This is a 12-item scale to assesses mistrust of mainstream health care systems and health care professionals by respondents ethnic or racial group. The responses included a Likert scale (*1-strongly disagree* to *5-strongly agree*) and a score range of 12-60 (18). Sample questions include people of my ethnic group cannot trust doctors, people from my ethnic group should not confide in health care workers because it will be used against them and I have personally been treated poorly or unfairly by doctors or health care workers because of my ethnicity.

### Data analysis section

The distribution of participants according to age, gender, marital status, education, and income, insurance, ethnicity, length of time in the U.S., birthplace and citizenship status were calculated. Mean values and corresponding standard deviations were calculated for mental health knowledge, stigma behavior, helping seek behaviors scores were calculated according to ethnicity and whether the participants had low (<10) or high depression scores (>10). The distribution of participants according to depression and ethnicity, citizenship, age group, and gender were calculated. Finally, we calculated mean medical mistrust scores according to ethnicity, citizenship status, age category, education, and gender. P-values for differences in mean scores were calculated using ANOVA and Chi-square for categorical variables. All analysis was performed using SAS Version 9.3 (SAS Institute).

## Results

Among the sample, 59.8% of participants were 34 or younger. The majority of the sample identified as male (57.2%) and reported being married (63.9%). Among the participants, 32.3% had no college education. A large proportion of the sample (43.1%) reported a personal income less than $30,000. The majority of participants reported having public health insurance (55.39%). Participants identified as African American (75.46%), African immigrant (19.33%), and Afro-Caribbean immigrant (4.83%). More than half of the participants (56.88%) reported living in the United States for more than 10 years. Most (76.21%) participants had United States citizenship.

**Table 1.** Descriptive demographics and general percentages for the sample

|  | n | % |
|--|---|---|
| Age |  |  |
| 34 or younger | 161 | 59.85 |
| 35 or older | 108 | 40.15 |
| Gender |  |  |
| Male | 154 | 57.25 |
| Female | 115 | 42.75 |
| Marital status |  |  |
| Married | 172 | 63.94 |
| Unmarried, living with a romantic partner | 45 | 16.73 |
| Never married | 29 | 10.78 |
| Separated | 7 | 2.60 |
| Divorced | 15 | 5.58 |
| Widowed | 1 | 0.37 |
| Education |  |  |
| Less than high school | 5 | 1.86 |
| High school diploma / GED | 44 | 16.36 |
| Trade school/vocational school | 38 | 14.13 |
| Some college, no degree | 91 | 33.83 |
| 2 year college | 44 | 16.36 |
| 4-year college degree | 40 | 14.87 |
| Master's degree | 4 | 1.49 |
| Doctoral degree | 3 | 1.12 |
| Income |  |  |
| \$9,999 or less | 15 | 5.58 |
| \$10,000 to \$29,999 | 101 | 37.55 |
| \$30,000 to \$49,999 | 119 | 44.24 |
| \$50,000 or more | 34 | 12.64 |
| Insurance |  |  |
| None | 60 | 22.30 |
| Public Insurance | 149 | 55.39 |
| Private | 60 | 22.30 |
| Ethnicity |  |  |
| African | 52 | 19.33 |
| African American | 203 | 75.46 |
| Afro-Caribbean | 13 | 4.83 |
| Other | 1 | 0.37 |
| How long in U.S.? |  |  |
| Less than 2 years | 6 | 2.23 |
| 2 to 5 years | 37 | 13.75 |
| 6 to 10 years | 73 | 27.14 |
| More than 10 years | 154 | 56.88 |
| Citizenship status |  |  |
| Citizen | 205 | 76.21 |
| Non-citizen | 64 | 23.79 |

**Table 2.**
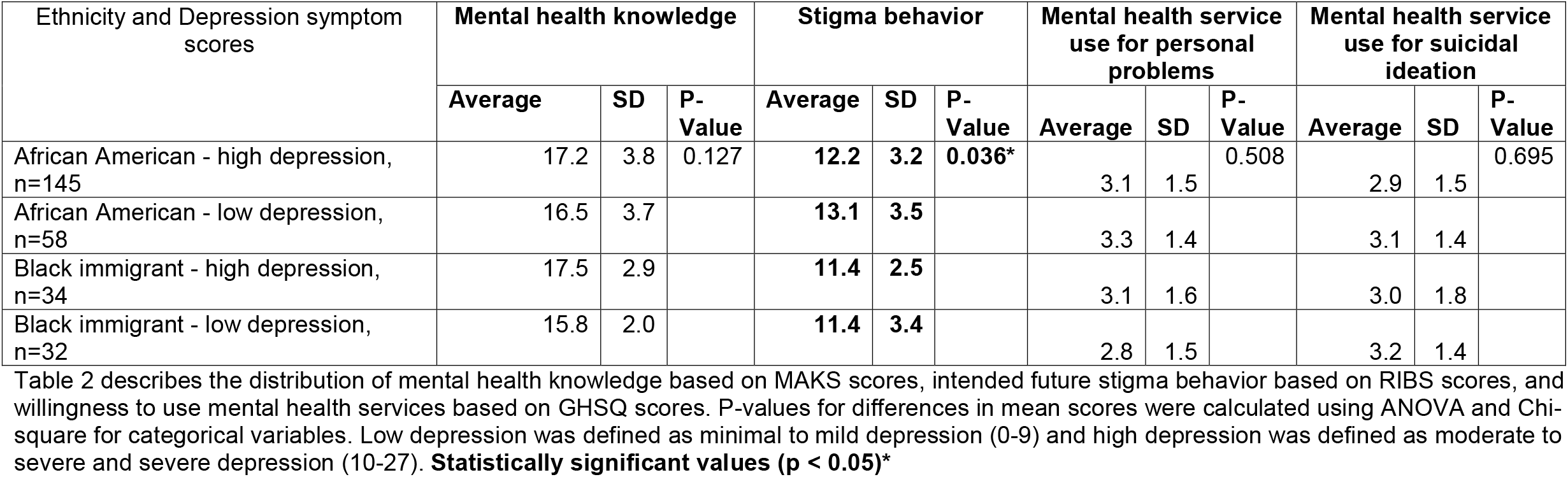
Distribution of averages of mental health knowledge, stigma behavior, willingness to seek help from mental health professional for personal and emotional problem and willingness to seek help from a mental health professional for suicidal ideation.

We assessed prevalence related to stigma behavior, higher social distance scores on the RIBS represent less stigmatizing behavior. African Americans who screened in the moderate to severe depression range had on average social distance scores that were lower (higher stigma) (mean = 12.2, SD = 3.2), hence greater stigma than African Americans who screened in the minimal to mild depression range (mean = 13.1, SD = 3.5). There was no difference in the mean value of stigma behavior between Black immigrants with moderate to severe depression screen and Black immigrants with minimal to mild depression (moderate to severe depression mean 11.4, SD = 2.5; minimal to mild depression mean 11.4, SD = 3.4). Black immigrants across the spectrum of depression scores had greater stigma than African Americans. African Americans with lower depression scores had the least amount of stigma in comparison across all four groups. These differences were statistically significant (p = 0.036).

**Figure 1.**
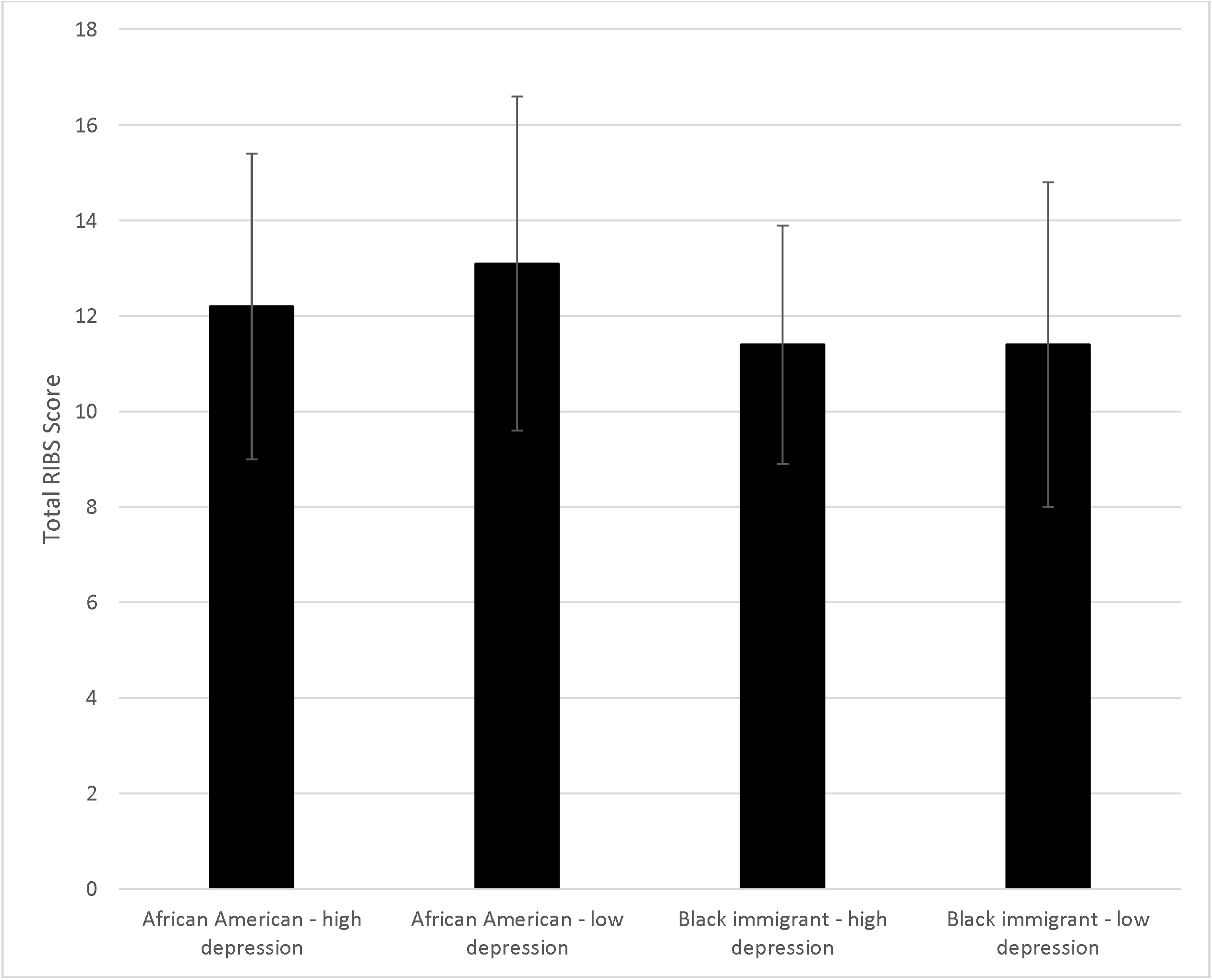
Stigma behavior (RIBS) scores according to ethnicity and depression symptom severity. Higher scores indicate less stigma behavior.

**Figure 4.**
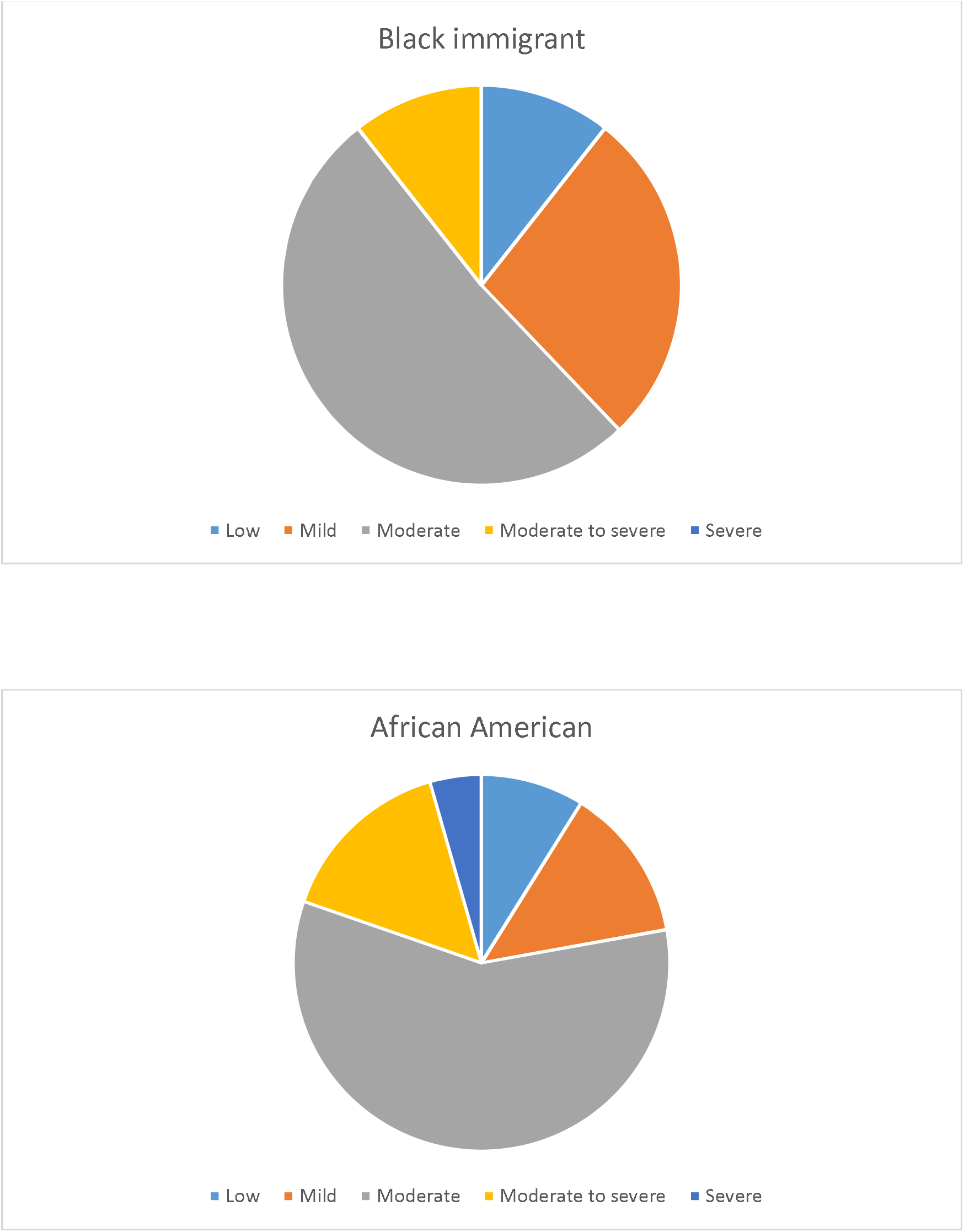
Depression symptom scores (low 0-4, mild 5-9, moderate 10-14, moderate to severe 15-19, severe 20-27) according to ethnicity

**Table 3.** Depression symptom status according to ethnicity, citizenship, age group, and gender

|  | **Minimal** | **Mild** | **Moderate** | **Moderate to severe** | **Severe** | **Total** | **P-value** |
| --- | --- | --- | --- | --- | --- | --- | --- |
| **African American?** | **n (%)** |  |  |  |  |  |  |
| Black immigrants | 7<br>(10.61) | 18<br>(27.27) | 34<br>(51.52) | 7 (10.61) | 0 (0.00) | 66 | 0.085 |
| African American | 18<br>(8.87) | 27<br>(13.3) | 118<br>(58.13) | 31<br>(15.27) | 9 (4.42) | 203 |  |
| **Citizen?** |  |  |  |  |  |  |  |
| Yes | 25<br>(11.71) | 33<br>(16.10) | 116<br>(56.59) | 27<br>(13.17) | 5 (2.44) | 205 | 0.072 |
| No | 1 (1.56) | 12<br>(18.75) | 36<br>(56.25) | 11<br>(17.19) | 4 (6.25) | 64 |  |
| **Age** |  |  |  |  |  |  |  |
| <35 | 10<br>(6.21) | 29<br>(18.01) | 89<br>(55.28) | 25<br>(15.53) | 8 (4.97) | 161 | 0.081 |
| >34 | 15<br>(13.89) | 16<br>(14.81) | 63<br>(58.33) | 13<br>(12.04) | 1 (0.93) | 108 |  |
| **Gender** |  |  |  |  |  |  |  |
| **Male** | **19**<br>**(12.34)** | **31**<br>**(20.13)** | **82**<br>**(53.25)** | **20**<br>**(12.99)** | **2 (1.3)** | **154** | **0.021\*** |
| **Female** | **6 (5.22)** | **14**<br>**(12.17)** | **70**<br>**(60.87)** | **18**<br>**(15.65)** | **7 (6.09)** | **115** |  |
| Total | 25 | 45 | 152 | 38 | 9 | 269 |  |

Table 3 shows depression symptom severity across ethnicity, citizenship, age group and gender. Depression symptom scores are shown (low 0-4, mild 5-9, moderate 10-14, moderate to severe 15-19, severe 20-27) along with prevalence in sample and percent in each group. P-values for differences in mean scores were calculated using ANOVA and Chi-square for categorical variables. **Statistically significant values ( $p < 0.05$ )\***

We assessed depression symptom severity according to gender. Male gender had a prevalence that was 1.7 times higher than female gender for mild depression (5-9). Female gender had a prevalence that was 1.2 times higher for moderate to severe depression (10-19) and 4.7 times higher for severe depression (20-27) compared to male gender. Differences in the distribution of depression levels by gender were statistically significant (p = 0.021).

**Table 4.** Medical mistrust according to ethnicity, citizenship status, age category, education, and gender

| **Ethnicity** | **Mean** | **SD** | **Min** | **Max** | **P-value** |
| --- | --- | --- | --- | --- | --- |
| Black immigrant | 2.87 | 0.46 | 1.67 | 3.83 | 0.108 |
| African American | 2.97 | 0.43 | 2.08 | 4.67 |  |
| **Citizen?** |  |  |  |  |  |
| **Yes** | **2.91** | **0.44** | **1.67** | **4.67** | **0.011\*** |
| **No** | **3.07** | **0.43** | **2.00** | **3.83** |  |
| **Age category** |  |  |  |  |  |
| <35 | 2.92 | 0.41 | 1.92 | 3.83 | 0.206 |
| >34 | 2.99 | 0.49 | 1.67 | 4.67 |  |
| **Education** |  |  |  |  |  |
| High school or less | 3.00 | 0.41 | 2.00 | 3.83 | 0.366 |
| Some college or more | 2.92 | 0.46 | 1.67 | 4.67 |  |
| **Gender** |  |  |  |  |  |
| Male | 2.95 | 0.47 | 1.92 | 3.83 | 0.856 |
| Female | 2.94 | 0.41 | 1.67 | 4.67 |  |

Table 4 shows medical mistrust scores across ethnicity, citizenship, age group and gender. Higher scores indicate greater mistrust. P-values for differences in mean scores were calculated using ANOVA and Chi-square for categorical variables. **Statistically significant values ( $p < 0.05$ )\***

We found that non-United States citizens reported higher medical mistrust (mean value difference = 0.16) compared to United States citizens and this difference was statistically significant (p = 0.011). There was negligible difference in medical mistrust across ethnicity, age, education and gender. African Americans reported slightly higher discriminatory experiences (mean value difference = 0.10) compared to Black immigrants. Older Black adults reported slightly higher medical mistrust (mean value difference = 0.07) compared to younger Black adults. Male gender reported slightly higher medical mistrust than female gender (mean value difference = 0.01).

## Discussion

This is the first study to date that fills a critical gap in the mental health research literature around the heterogeneity among Black adults across social determinants of mental health related to ethnicity, depression symptom severity, mental health knowledge, stigma behavior, help seeking behavior and medical mistrust. Assessing the heterogeneous characteristics within the Black population provides increased understanding of the diversity within the community and strengthens the effectiveness of mental health interventions to address disparities and unmet needs among Black people (2, 19). This enhances innovation in modeling our mental health systems to meet the needs of Black people in more specific and effective ways towards closing public health gaps in Black mental health care (20, 21).

Our study highlights the critical need to consider the multiple identities among Black adults in addressing mental health system experiences and needs (2, 12, 22). Intersectionality theory applied to health refers to understanding the role of social context and characteristics in explaining health inequities by highlighting identity across race, age, gender, sexuality, ethnicity and other identities (12, 13). We saw variations in demographic characteristics across ethnicity, citizenship, age and gender. In this study, African Americans reported twice the level of moderate to severe depressive symptoms compared to Black immigrants, whereas, Black immigrants reported twice the level of minimal to mild depressive symptoms. Previous studies are more general in using prevalence data (2, 23), reporting on overall outcomes, rather than differences in symptom severity. Male adults reported a level of mild depression symptoms that was 1.7 times higher than female Black adults, whereas, female Black adults reported severe depression that was 4.7 times higher than male Black adults. Specific differences in symptom severity is critical to understand as it may influence the timing of presentation for mental health treatment as well as the course of illness and management of depression across ethnicity and gender.

Intersectional identities among Black adults across ethnicity, citizenship status, age, and gender influences experiences of discrimination within the mainstream health system (11-13). African Americans report marginally higher mistrust than Black immigrants whereas those who were not United States citizens reported higher mistrust than US citizens. Older Black adults reported greater mistrust than younger Black adults. Male respondents reported greater mistrust than female respondents. The variations in ethnicity, citizenship, age and gender in experiences of medical mistrust is evidence of the ways in which addressing mental health needs in a homogenous manner in the Black community leads to missing how various identities impact mental health experiences. For example, among non-United citizens, stressors around acculturation in relation to migration may lead to greater mistrust of mainstream health systems (24, 25). Previous studies reveal age and generational status has been shown to influence interpretation of discriminatory experiences (26, 27), and our study highlights the differences in medical mistrust based on specific characteristics among Black adults in our sample.

African Americans with higher depressive symptoms had greater stigma than African Americans with lower depressive symptoms. There was no difference between Black immigrants with higher depressive symptoms compared to lower depressive symptoms, though Black immigrants reported higher stigma behavior than African Americans. Stigma is a known barrier to accessing mental health services and therefore it is critical to address stigma (6, 28, 29), especially in the context of greater illness severity and delay in treatment seeking. Our study shows the importance of considering variations in stigma behavior across ethnicity and illness severity in order to address mental health needs using approaches that do not treat Black adults as a monolithic group in their mental health needs.

### Study Limitations

While this was a cross-sectional study, given the current limited data in this area of research, the purpose of the study was successful in showing evidence that there is a critical need to understanding the heterogeneity within the Black adult population in order to address unmet mental health needs. While we compared African Americans to Black immigrants in this study with a sample size of 269 adults, future studies would be able to further assess the intersectionality framework by comparing overlapping identities such as across age, ethnicity, citizenship and migrant status collectively rather than independent comparisons. We lay a strong foundation for the need to go beyond assessment of mental health outcomes such as prevalence of depression, but also consider variations across symptom severity and psychosocial factors related to the social determinants of mental health. As a descriptive study, we sought to show mean differences across demographic characteristics and depression severity, future studies would benefit from assessing objective behavior (such as actual use of mental health services within health system records).

## Conclusion

Our study shows heterogeneity across ethnicity, citizenship, age, gender and depression symptom severity among Black adults. We provide data on differences in this population using an intersectionality framework and examining social determinants of mental health. There were differences in stigma behavior across ethnicity and depression severity. There were also differences in medical mistrust based on citizenship status with non-United States citizens reporting greater medical mistrust. The distribution of depression severity varied across gender and has implications on our approach to clinical care across gender identities in the Black population. Ongoing research in this area will contribute to closing the mental health gap experienced by Black adults within our current health systems that tends to pay little attention to the intersectional identities of Black adults that determine mental health outcomes.

## Data Availability

All data produced in the present work are contained in the manuscript

